# Inter-population differences of allele frequency and regulome tagging are associated with the heterogeneity of loci identified by cross-ancestry genome-wide association studies

**DOI:** 10.1101/2020.09.13.20193656

**Authors:** Antonella De Lillo, Salvatore D’Antona, Gita A. Pathak, Frank R. Wendt, Flavio De Angelis, Maria Fuciarelli, Renato Polimanti

## Abstract

To investigate cross-ancestry genetics of complex traits, we conducted a phenome-wide analysis of loci with heterogeneous effects across African, Admixed-American, Central/South Asian, East Asian, European, and Middle Eastern participants of UK Biobank (N=441,331). Testing 843 phenotypes, we identified 82 independent genomic regions mapping variants showing genome-wide significant (GWS) associations (p<5×10^−8^) in the trans-ancestry meta-analysis and GWS heterogeneity among the ancestry-specific effects. These included: i) loci with GWS association in one ancestry and concordant but heterogeneous effects among the other ancestries; ii) loci with a GWS association in one ancestry group and an experiment-wide significant discordant effect (p<6.1×10^−4^) in at least another ancestry. Since the trans-ancestry GWS associations were mostly driven by the European-ancestry sample size, we investigated the differences of allele frequency (Δ_AF_) and linkage-disequilibrium regulome tagging (Δ_LD_) between European populations and the other ancestries. Within loci with concordant effects, the degree of heterogeneity was associated with European – Middle Eastern Δ_AF_ (p=9.04×10^−6^) and Δ_LD_ of European populations with respect to African, Admixed-American, and Central/South Asian groups (p=8.21×10^−4^, p=7.17×10^−4^, and p=2.16×10^−3^, respectively). Within loci with discordant effects, Δ_AF_ and Δ_LD_ of European populations with respect to African and Central/South Asian ancestries was associated with the degree of heterogeneity (Δ_AF_: p=7.69×10^−3^ and p=5.31×10^−3^, Δ_LD_: p=0.016 and p=2.65×10^−4^, respectively). Considering the traits associated with cross-ancestry heterogeneous loci, we observed enrichments for blood biomarkers (p=5.7×10^−35^) and physical appearance (p=1.38×10^−4^). This suggests that these specific phenotypic domains may present considerable cross-ancestry heterogeneity due to large allele frequency and LD variation among worldwide populations.

## Introduction

Genome-wide association studies (GWAS) are a powerful tool to identify genetic variants associated with human traits and diseases (1). As of December 15, 2020, 4,809 publications and 227,262 associations have been listed in the GWAS Catalog (2). This unprecedented amount of information has revolutionized our understanding of the predisposition to complex phenotypes, demonstrating that a large portion of the heritability of complex traits resides in common genetic variation (i.e., polymorphisms in the human genome that show a minor allele frequency (MAF) greater than 1%) (1). In recent years, the investigations of massive cohorts from 100,000 to more than 1,000,000 participants were possible because of large collaborative projects combining numerous studies (3-6), the availability of biobanks enrolling an unprecedented number of participant (7-9), and collaboration with direct-to-consumer genetic testing companies (10). These large-scale GWAS identifying ever-greater numbers of risk loci with ever-smaller individual effects demonstrated that the genetic architecture of common diseases is highly polygenic and their heritability is likely due to the contribution of several thousand (or even more) risk loci across the human genome (11-14). One of the main GWAS promises is that the knowledge gained can be used to develop genetic instruments useful to predict disease risk, treatment response, and disease prognosis. Leveraging data generated by large-scale GWAS, a growing number of studies are developing approaches to test the utility of polygenic information with respect to the human phenotypic spectrum (15-18). Although these successful experiments strongly support the movement towards the application of GWAS data to develop new strategies to prevent and treat human diseases, important challenges remain. Among them, one of the most pressing is related to the limited ancestry and ethnic diversity of large-scale GWAS that have created a large gap between the genetic data available for populations of European vs. non-European descent (19). Applying GWAS data generated from European-ancestry cohorts to non-European individuals raises serious issues, including much lower predictive power than that observed in comparisons between like populations (20, 21) and possible biases (e.g., reflecting unaccounted population stratification rather than the phenotype of interest) due to the genetic variability among human populations (22, 23). The most reliable solution to this problem is to conduct large-scale GWAS in populations with non-European ancestry. Ongoing efforts such as the Million Veteran Program (24) and the AllofUS Research Program (25) are investigating multiple ancestry groups representative of the US population to reduce this gap. Although these projects are expected to reduce the population disparities in human genetic research, this is likely to be a long-term outcome. To date, to contribute to a more comprehensive understanding of human genetic diversity, we can leverage the data available, combining large-scale genome-wide association datasets generated from cohorts, mainly including participants of European descent with reference panels representative of the genetic diversity among worldwide populations (26-29). In the present study, we focused our attention on the UK Biobank (UKB). This large cohort includes more than 500,000 participants with >90% of them as British individuals of European descent (30). In addition to participants of European descent, the UKB cohort includes individuals of African (AFR), Admixed-American (AMR), Central/South Asian (CSA), East Asian (EAS), and Middle Eastern (MID) ancestral background. Combining UK Biobank cross-ancestry data with information regarding the inter-population variability in the linkage-disequilibrium (LD) tagging of regulatory elements, we investigated the cross-ancestry heterogeneity of loci associated with the human phenotypic spectrum.

## Materials and Methods

### UK Biobank

UKB is a large population-based prospective study to explore different life-threatening disorders using information about environmental factors and genes in order to improve diagnosis and treatment (9). A wide variety of phenotypic information, including socio-demographic and lifestyle factors, electronic health records data, and physiological conditions have been collected for more than 500,000 UKB participants (30). UKB genetic data were used to generate genome-wide association datasets that can be employed to explore the genetics of complex traits. In our study, we used the Pan-UKB genome-wide association statistics generated from the analysis of six ancestries (AFR N=6,636; AMR N=980; CSA N=8,876; EAS N=2,709; EUR N 429,531; MID N=1,599). Pan-UKB data are available at https://pan.ukbb.broadinstitute.org/downloads. A detailed description of the methods used to generate these data is available at https://pan.ukbb.broadinstitute.org/. Briefly, the ancestry assignment of UKB participants was conducted with respect to combined reference data from the 1000 Genomes Project (1KG) (31) and the Human Genome Diversity Project (HGDP) (32) using a two-stage approach: i) assign continental ancestries and ii) prune ancestry outliers within continental groups. The top 6 principal components (PCs) from the reference data were used to train a random forest classifier that was then applied to the UK Biobank PC data. UKB participants were assigned to an ancestry group based on a random forest probability >50%. Individuals with a probability of <50% were excluded from the analysis. The genetic association analysis investigated variants with an imputation INFO score > 0.8 and a minimum allele count of 20. Phenotypes analyzed included binary, ordinal, and continuous traits. For binary traits, a minimum of 50 cases were required in each ancestry group with the exception of the European-ancestry sample where at least 100 cases were required. To investigate cross-ancestry heterogeneity, we analyzed 843 traits (Supplemental Table 1) assessed across all six ancestry groups available in the UKB cohort. Within each ancestry, the genome-wide association analysis was conducted using the SAIGE (Scalable and Accurate Implementation of GEneralized) mixed model (33) and including a kinship matrix as a random effect and covariates as fixed effects. The covariates included age, sex, age×sex, age^2^, age^2^×sex, and the top-10 within-ancestry principal components. The ancestry-specific genome-wide associations were meta-analyzed using the inverse variance weighted method and a heterogeneity test of the cross-ancestry meta-analysis was also performed.

### Regulome LD tagging properties across ancestry groups

To investigate how the LD structure across worldwide populations affects the ability of the GWAS index variants to tag functional elements in the surrounding regions, we leveraged 1KG reference superpopulations (31) and information regarding regulatory variants from RegulomeDB (34). We used LD information from the 1KG reference panel instead of the UKB cohort, because 1KG data generated from whole-genome sequencing are more informative of LD tagging variability than UKB data generated from the GWAS array plus imputation. Using LDlink (35, 36), we tested the effect of the LD structure variability across ancestry groups on the ability of variants to tag (measured as LD R^2^) functional variants in the surrounding regions (±500Kb). The 1KG reference superpopulations include AFR, AMR, EAS, EUR, and South Asian. UKB CSA sample was defined by combining 1KG South Asian and HGDP South/Central Asian reference data. Accordingly, we refer to the 1KG South Asian reference panel as CSA hereafter. RegulomeDB (34) was used to score the regulatory effect of the tagged variants on the basis of high-throughput, experimental data sets as well as computational predictions and manual annotations. RegulomeDB scoring scheme ranges from 1 (highest number of known and predicted data regarding regulatory function) to 7 (lowest number of known and predicted data regarding regulatory function). To quantify the ancestry-specific ability of the GWAS index variants to tag regulatory elements in their surrounding regions, we defined an LD regulome-tagging score (LD-RTS) for each GWAS index variant calculated as: 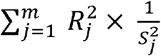, where *j* is a genetic polymorphism in *m* polymorphisms within ±500Kb of the GWAS index variant, *R* is the LD correlation coefficient between the polymorphism *j* and the GWAS index variant, *S* is the RegulomeDB score of the polymorphism *j*. LD-RTS scores were calculated for each GWAS index variants with respect to the LD information available from each of the 1KG reference superpopulations.

### Non-parametric regression analyses

To understand the relationship between the heterogeneity observed in the trans-ancestry meta-analysis and genetic variation among worldwide populations, we conducted non-parametric regression analyses. We decided to apply non-parametric tests, because of the non-normal distribution of the variables investigated. Because of the much larger UKB EUR sample, the trans-ancestry GWS associations were mostly driven by EUR-specific results. Accordingly, the heterogeneity observed in the UKB trans-ancestry meta-analysis should be mainly due to genetic differences of EUR populations with respect to the other ancestry groups. For this reason, we considered the differences of the allele frequency and LD-RTS (Δ_AF_ and Δ_LD_, respectively) of European populations with respect to the other ancestries investigated. These variables were entered in the non-parametric regression models considering the degree of heterogeneity as the outcome of interest. The latter was quantified as the negative of the base-10 logarithm of the p-values obtained from the trans-ancestry meta-analysis heterogeneity test. We applied a two-stage approach. Initially, we applied two-dimensional linear models to investigate the relationship between the degree of heterogeneity and each of the Δ_AF_ and Δ_LD_ calculated using median-based linear regression (MBLM) approach (available at https://cran.r-project.org/web/packages/mblm/index.html). Then, we entered the significant MBLM variables in a multi-dimensional regression applying the locally-estimated scatterplot smoothing (LOESS) process (available at https://www.rdocumentation.org/packages/stats/versions/3.6.2/topics/loess) and testing it using the generalized additive model (GAM) approach (available at https://cran.r-project.org/web/packages/gam/index.html).

### Enrichment analysis for phenotypic domains

To test whether there is an over-representation of certain phenotypic domains among traits associated with loci presenting cross-ancestry heterogeneity, we calculated the significance of the phenotypic enrichment applying the cumulative distribution function of the hyper-geometric distribution to the proportions of the phenotypic domains associated with the GWAS index variants investigated with respect to the proportion of phenotypic domains across the overall phenotypic spectrum investigated (Supplemental Table 1).

## Results

Considering Pan-UKB genome-wide association statistics related to 843 phenotypes investigated across the six ancestry groups available, we identified 20,287 variants presenting genome-wide significance (GWS, p<5×10^−8^) in both the trans-ancestry meta-analysis and the heterogeneity test (Supplemental Table 2). To identify the number of independent signals, we decided to apply a conservative approach based on positional mapping of this variant list. We did not use LD information, because the cross-ancestry meta-analysis was generated from six ancestry groups with different LD structure. Accordingly, we defined independent blocks considering a pairwise distance of 10Mb. Specifically, we applied a two-variant window and assigned variants to the same block if closer than 10Mb; a novel block is defined when two variants are more distant than 10 Mb. Following this criterion, we identified 82 independent genomic regions mapping the 20,287 variants presenting GWS trans-ancestry associations and GWS heterogeneity among the ancestry-specific effects. Within each block, a GWAS index variant was defined based on the significance of the cross-ancestry heterogeneity test (Supplemental Table 3). Among them, we observed two different scenarios: i) loci with GWS association in one ancestry and concordant but heterogeneous effects among the other ancestries (i.e., cross-ancestry differences in the effect size); ii) loci with a GWS association in one ancestry group and an experiment-wide significant discordant effect (p<6.1×10^−4^, i.e., Bonferroni correction accounting for the number of independent genomic regions) in at least one other ancestry. Among the concordant but heterogeneous index variants, the strongest degree of heterogeneity was observed in the association of *SLC45A2* rs35390*A with “Hair color (natural, before greying): black” (trans-ancestry beta=-0.456, p=1.95×10^−78^, heterogeneity p=6.49×10^−192^) that showed a much larger effect size in UKB EUR participants (EUR beta=-1.928, p=5.12×10^−265^) than in other ancestries (AMR beta=-0.629, p=1.03×10^−5^; MID beta=-0.380, p=1.54×10^−4^). With respect to the “discordant-effect” loci, the association of *LUC7L* rs7189975*A with mean corpuscular haemoglobin (trans-ancestry beta=0.105, p=2.82×10^−102^, heterogeneity p=6.03×10^−104^) was positive in the UKB EUR participants (EUR beta=0.134, p=2.26×10^−151^) and negative in the UKB AFR participants (AFR beta=-0.336, p=1.81×10^−58^). Due to the large sample size of the UKB EUR sample, most of the trans-ancestry associations of the GWAS index variants were driven by the EUR-specific analysis. Only three trans-ancestry associations presented the strongest ancestry-specific signals in non-EUR samples: *UGT1* family members A3-A10 rs12466997*C (total bilirubin; trans-ancestry beta=-0.107, p=3.54×10^−91^; AFR beta=-0.554, p=5.18×10^−142^; heterogeneity p=1.5×10^−109^), *F8* rs782604098*ATG (glycated haemoglobin; trans-ancestry beta=-0.027, p=2.71×10^−9^; AFR beta=-0.544, p=1.45×10^−109^; heterogeneity p=5.59×10^−103^), and *ABO* rs9411476*A (alkaline phosphatase; trans-ancestry beta=-0.088, p=2.99×10^−13^; AFR beta=-0.228, p=3.21×10^−16^; heterogeneity p=5×10^−24^). The heterogeneous effect of these AFR-driven associations showed concordant directions across the other ancestries. Table 1 reports the details of the associations described above.

**Table 1:**
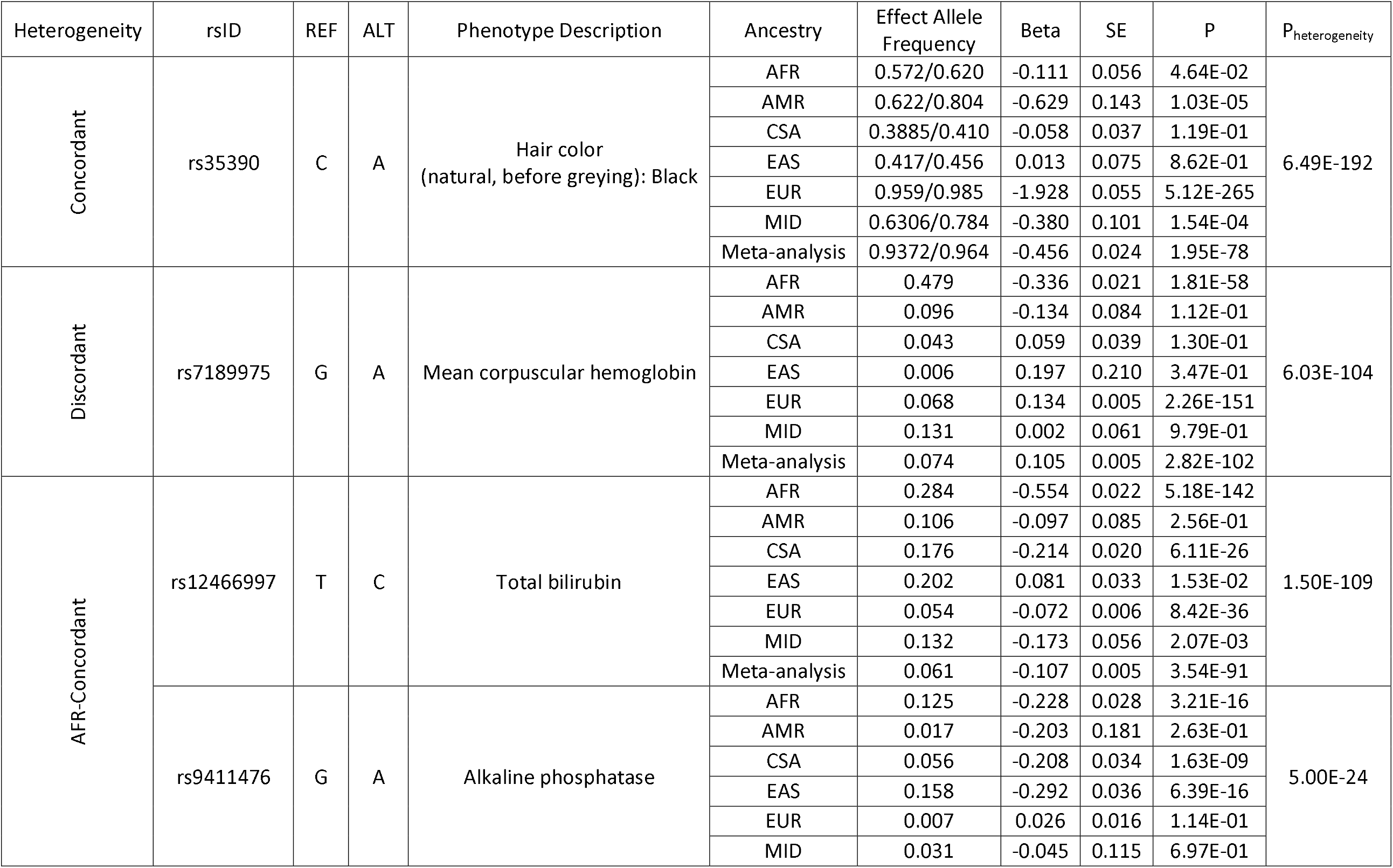
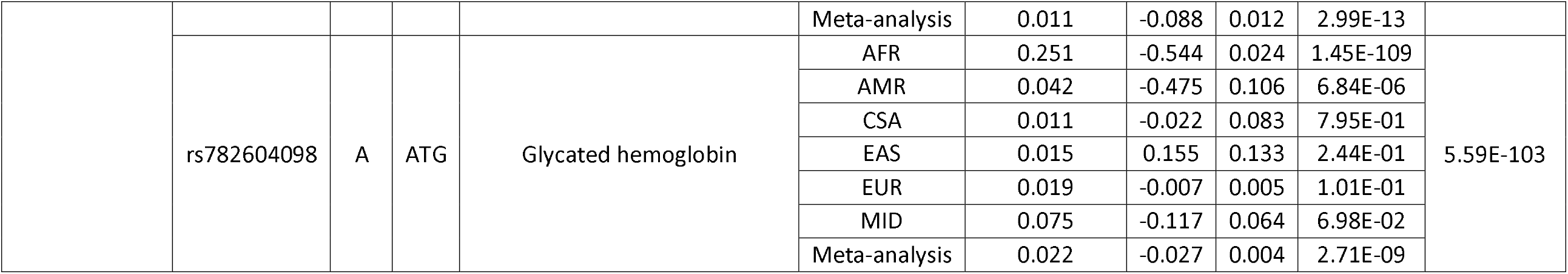
Loci with most significant concordant and discordant heterogeneity and loci with cross-ancestry associations driven by the UKB AFR sample. The alternate allele (ALT) is the effect allele. For binary outcomes, we report the effect allele frequency for both cases and controls.

| Heterogeneity | rsID | REF | ALT | Phenotype Description | Ancestry | Effect Allele Frequency | Beta | SE | P | P<sub>heterogeneity</sub> |
| --- | --- | --- | --- | --- | --- | --- | --- | --- | --- | --- |
| Concordant | rs35390 | C | A | Hair color<br>(natural, before greying): Black | AFR | 0.572/0.620 | -0.111 | 0.056 | 4.64E-02 | 6.49E-192 |
|  |  |  |  |  | AMR | 0.622/0.804 | -0.629 | 0.143 | 1.03E-05 |  |
|  |  |  |  |  | CSA | 0.3885/0.410 | -0.058 | 0.037 | 1.19E-01 |  |
|  |  |  |  |  | EAS | 0.417/0.456 | 0.013 | 0.075 | 8.62E-01 |  |
|  |  |  |  |  | EUR | 0.959/0.985 | -1.928 | 0.055 | 5.12E-265 |  |
|  |  |  |  |  | MID | 0.6306/0.784 | -0.380 | 0.101 | 1.54E-04 |  |
|  |  |  |  |  | Meta-analysis | 0.9372/0.964 | -0.456 | 0.024 | 1.95E-78 |  |
| Discordant | rs7189975 | G | A | Mean corpuscular hemoglobin | AFR | 0.479 | -0.336 | 0.021 | 1.81E-58 | 6.03E-104 |
|  |  |  |  |  | AMR | 0.096 | -0.134 | 0.084 | 1.12E-01 |  |
|  |  |  |  |  | CSA | 0.043 | 0.059 | 0.039 | 1.30E-01 |  |
|  |  |  |  |  | EAS | 0.006 | 0.197 | 0.210 | 3.47E-01 |  |
|  |  |  |  |  | EUR | 0.068 | 0.134 | 0.005 | 2.26E-151 |  |
|  |  |  |  |  | MID | 0.131 | 0.002 | 0.061 | 9.79E-01 |  |
|  |  |  |  |  | Meta-analysis | 0.074 | 0.105 | 0.005 | 2.82E-102 |  |
| AFR-Concordant | rs12466997 | T | C | Total bilirubin | AFR | 0.284 | -0.554 | 0.022 | 5.18E-142 | 1.50E-109 |
|  |  |  |  |  | AMR | 0.106 | -0.097 | 0.085 | 2.56E-01 |  |
|  |  |  |  |  | CSA | 0.176 | -0.214 | 0.020 | 6.11E-26 |  |
|  |  |  |  |  | EAS | 0.202 | 0.081 | 0.033 | 1.53E-02 |  |
|  |  |  |  |  | EUR | 0.054 | -0.072 | 0.006 | 8.42E-36 |  |
|  |  |  |  |  | MID | 0.132 | -0.173 | 0.056 | 2.07E-03 |  |
|  |  |  |  |  | Meta-analysis | 0.061 | -0.107 | 0.005 | 3.54E-91 |  |
|  | rs9411476 | G | A | Alkaline phosphatase | AFR | 0.125 | -0.228 | 0.028 | 3.21E-16 | 5.00E-24 |
|  |  |  |  |  | AMR | 0.017 | -0.203 | 0.181 | 2.63E-01 |  |
|  |  |  |  |  | CSA | 0.056 | -0.208 | 0.034 | 1.63E-09 |  |
|  |  |  |  |  | EAS | 0.158 | -0.292 | 0.036 | 6.39E-16 |  |
|  |  |  |  |  | EUR | 0.007 | 0.026 | 0.016 | 1.14E-01 |  |
|  |  |  |  |  | MID | 0.031 | -0.045 | 0.115 | 6.97E-01 |  |
|  |  |  |  |  | Meta-analysis | 0.011 | -0.088 | 0.012 | 2.99E-13 |  |
|  | rs782604098 | A | ATG | Glycated hemoglobin | AFR | 0.251 | -0.544 | 0.024 | 1.45E-109 | 5.59E-103 |
|  |  |  |  |  | AMR | 0.042 | -0.475 | 0.106 | 6.84E-06 |  |
|  |  |  |  |  | CSA | 0.011 | -0.022 | 0.083 | 7.95E-01 |  |
|  |  |  |  |  | EAS | 0.015 | 0.155 | 0.133 | 2.44E-01 |  |
|  |  |  |  |  | EUR | 0.019 | -0.007 | 0.005 | 1.01E-01 |  |
|  |  |  |  |  | MID | 0.075 | -0.117 | 0.064 | 6.98E-02 |  |
|  |  |  |  |  | Meta-analysis | 0.022 | -0.027 | 0.004 | 2.71E-09 |  |

To understand how the genetic variation across ancestries affects the heterogeneity observed among the ancestry-specific associations, we investigated differences with respect to allele frequency and linkage disequilibrium. Figure 1 reports Spearman’s correlation among the allele frequencies and LD-RTS of the 82 GWAS index variants among the ancestries investigated. Since EUR sample was the main driver of most of the trans-ancestry associations investigated (96%), we analyzed the relationship of the degree of heterogeneity among these associations with respect to the Δ_AF_ and Δ_LD_ of the European populations with the other ancestry groups (Table 2). Among the concordant but heterogeneous associations, the MBLM two-dimensional analysis highlighted the effect of four variables on the degree of heterogeneity: the EUR-MID Δ_AF_ (p_MBLM_ =9.04×10^−6^) and the Δ_LD_ of EUR sample with respect to AFR (p_MBLM_=8.21×10^−3^), AMR (p_MBLM_=7.17×10^−4^), and CSA (p_MBLM_=2.16×10^−3^). The LOESS-GAM multi-dimensional analysis confirmed that the effect of EUR-MID Δ_AF_ (p_LOESS-GAM_=3.76×10^−7^), EUR-AFR Δ_LD_ (p_LOESS-GAM_=0.026), and EUR-AMR Δ_LD_ (p_LOESS-GAM_=3.47×10^−3^) are independent of each other. A different pattern was observed with respect to the GWAS index variants with discordant, heterogeneous effects. Among them, the degree of heterogeneity observed was associated with the differences of EUR populations with AFR and CSA ancestries (EUR-AFR: Δ_AF_ p_MBLM_=7.69×10^−3^, Δ_LD_ p_MBLM_=0.016; EUR-CSA: Δ_AF_ p_MBLM_=5.31×10^−3^, Δ_LD_ p_MBLM_=5.31×10^−3^). However, these effects appeared to be driven by EUR-AFR Δ_AF_ (p_LOESS-GAM_=0.026). To explore further the effect of Δ_AF_ and Δ_LD_ on cross-ancestry heterogeneity, we analyzed their associations considering all GWAS index variants (Supplemental Table 4). Across loci with concordant and discordant heterogeneous cross-ancestry effects, the degree of heterogeneity was associated independently with EUR-MID Δ_AF_ (p_LOESS-GAM_=8×10^−6^), EUR-AFR Δ_LD_ (p_LOESS-GAM_=5.1×10^−3^), and EUR-AMR (p_LOESS-GAM_=4.4×10^−4^).

**Table 2:**
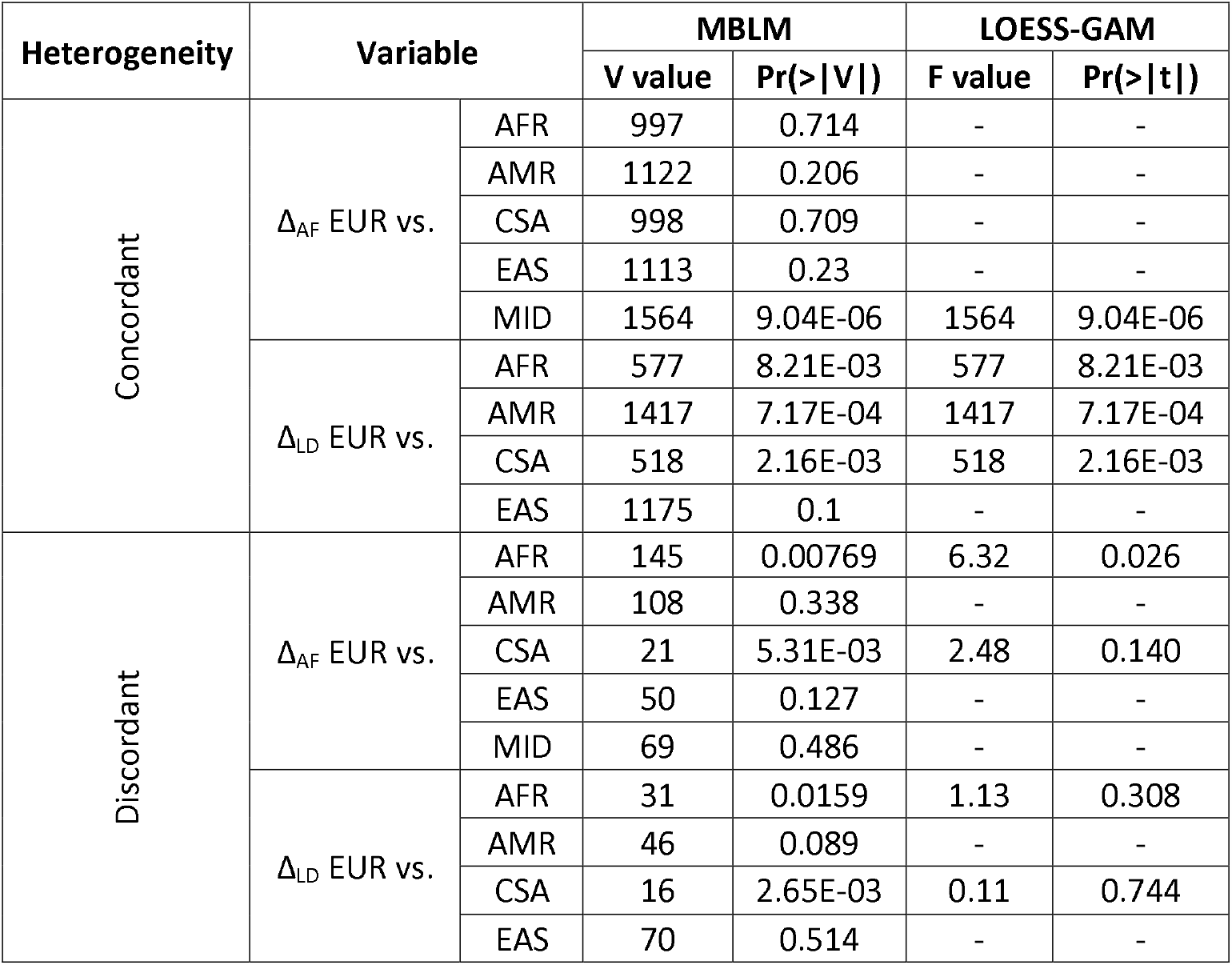
Association of Δ_AF_ and Δ_LD_ with the degree of heterogeneity considering GWAS index variants with concordant and discordant heterogeneity.

**Figure 1:**
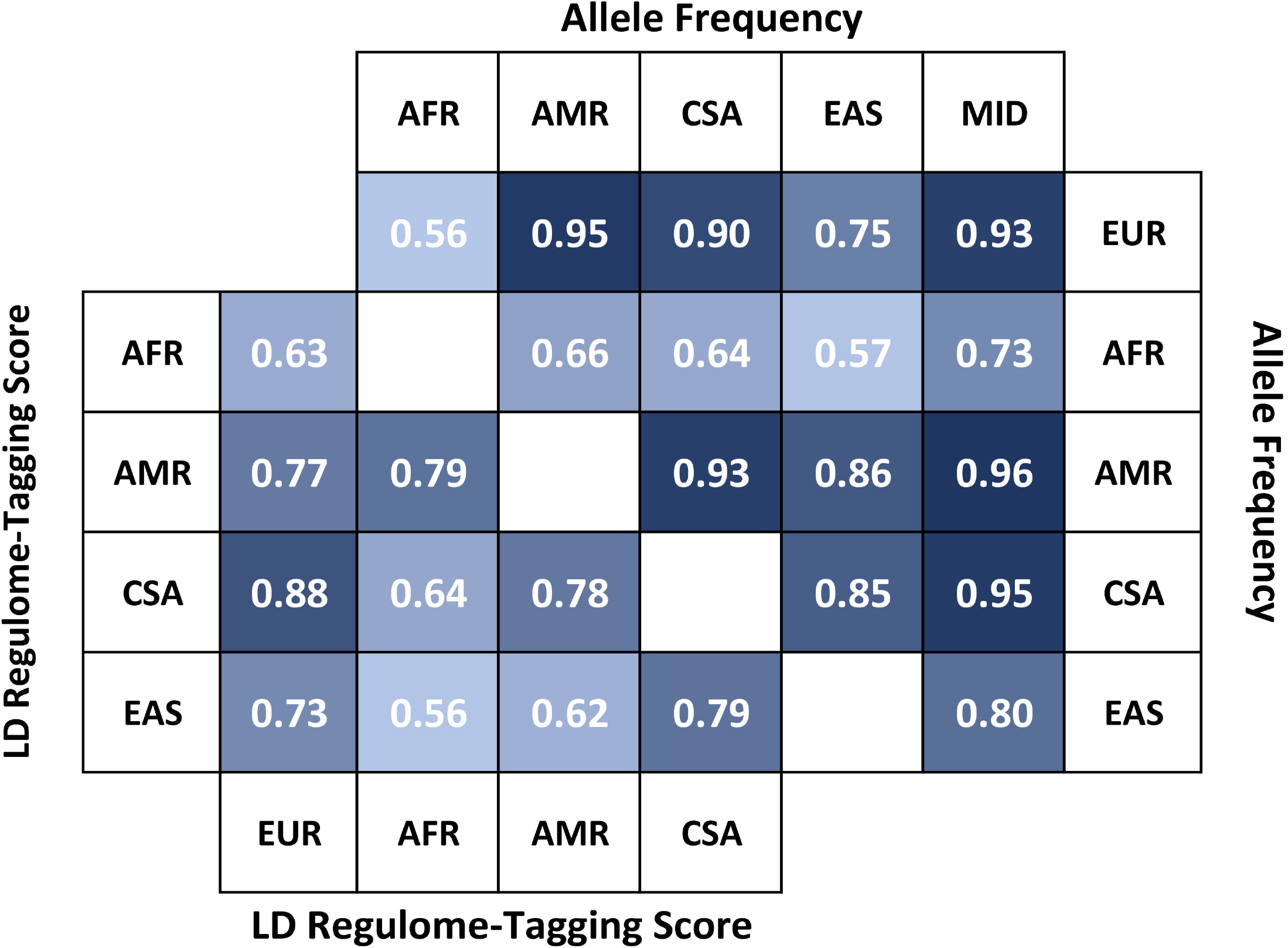
Correlation (Spearman’s rho) among allele frequencies (upper triangle) and LD regulome-tagging score (lower triangle) of the ancestral groups investigated considering the GWAS index variants investigated. The correlations presented survive Bonferroni multiple testing correction accounting for the number of tests performed.

The 82 GWAS index variants showed the strongest association with 40 traits belonging to three phenotypic domains: blood biomarkers (82.5%), physical appearance (10%), and anthropometric traits (7.5%). Accordingly, we observed that the same trait was associated with multiple GWAS index variants located in independent genomic regions. Among blood biomarkers, low-density lipoprotein (LDL) cholesterol adjusted by medication showed five independent associations with loci with cross-ancestry heterogeneous effects (i.e., *PCSK9* rs2479413 on chromosome 1, *DYNC2LI1* rs4953016 on chromosome 2, *ANKRD31* rs55810502 on chromosome 5, *TXNL4B* rs217181 on chromosome 16, and *SPC24* rs79668907 on chromosome 19). Similarly, “Hair color (natural, before greying): black” was associated with four independent loci with cross-ancestry heterogeneity (i.e., *SLC45A2* rs35390 on chromosome 5, *IRF4* rs11308001 on chromosome 6, rs11437447 on chromosome 12, and *SLC24A4* rs4904871 on chromosome 14). Other 20 traits in these phenotypic domains showed an association with at least two GWAS index variants. Comparing the distribution of phenotypic domains associated with loci with cross-ancestry heterogeneous effects with that of the 843 traits tested initially, we observed an over-representation for associations with blood biomarkers (enrichment=11.04, p=5.7×10^−35^) and traits related to physical appearance (enrichment=12.04, p=1.38×10^−4^).

## Discussion

To provide a more comprehensive understanding of the genetics of complex traits across worldwide populations, we investigated heterogeneity among loci identified by a cross-ancestry GWAS meta-analysis. The results obtained provide a comprehensive overview of how genetic differences among human population groups affect genetic associations of complex traits. Specifically, we observed that loci showing cross-ancestry heterogeneous effects present specific genetic and phenotypic characteristics.

Previous studies showed how allele frequency differences among human populations affect certain genotype-phenotype associations, because the number of effect-allele carriers changes the statistical power of the association analysis conducted (37). Similarly, the LD structure among human populations plays a key role in the functional implication of the variants identified by GWAS (38, 39). Indeed, it has been proposed that cross-ancestry meta-analyses can be a useful tool to fine map causal loci responsible for GWS loci (40-42). Our study demonstrates that both allele frequency and LD differences among human populations are significant contributors to the cross-ancestry heterogeneity across the human phenotypic spectrum. Considering the population diversity of the UKB cohort, we identified many loci with a cross-ancestry GWS association with a certain trait and a GWS heterogeneity among the ancestry-specific effects. The heterogeneity observed was both qualitative and quantitative. Among the GWAS index variants located in independent genomic regions, 16 loci (22%) showed qualitative heterogeneity among the ancestry specific effects, i.e., a GWS association in one ancestry group and an experiment-wide significant discordant effect in at least one another ancestry group. Applying a two-dimensional model (i.e., MBLM approach), the degree of heterogeneity was associated with the genetic differences of EUR ancestry with respect to AFR and CSA population groups. However, the multidimensional LOESS-GAM approach highlighted that the qualitative heterogeneity observed was primarily driven by the EUR-AFR Δ_AF_. This finding is in line with human evolutionary history. Because of the African origin of the human species and the subsequent out-of-Africa bottlenecks (43, 44), African populations present a much greater genetic variation than any other human group. Previous investigations highlighted how these genetic differences affect the statistical power of genetic association analyses (45). To our knowledge, the present findings represent the first comprehensive assessment of how the genetic diversity of African populations can lead to a discordant effect in genotype-phenotype associations when compared to other ancestries. A different scenario was observed with respect to the loci with quantitative heterogeneity (i.e., concordant effects with heterogeneous effect size). The degree of heterogeneity among these loci was independently associated with primarily EUR-MID Δ_AF_ and EUR-AMR Δ_LD_ and to a lesser extent with the EUR-AFR Δ_LD_. This highlights that, while the diversity of AFR populations is still a significant contributor, the cross-ancestry quantitative heterogeneity observed within the UK Biobank cohort was more strongly affected by differences of EUR ancestry with MID and AMR populations. This is particularly interesting because of the demographic history of these two human groups. Indeed, although they occurred on a different time scale and extent, a certain degree of admixture is present in both. AMR populations present an admixture of Native American, Sub-Saharan African, and European ancestries and the proportions of these three components can vary greatly across the American continent reflecting the pre-Colombian civilizations, the European colonization, and the Atlantic slave trade (46). MID populations present an admixture of Mediterranean African, Southern European, and Central Asian ancestries related to the demographic history that occurred over a long period of time in that region (e.g., rise and fall of empires, invasions, migrations, and trade) (47). Accordingly, our study highlights that the genetic admixture is likely a key mechanism contributing to the qualitative heterogeneity observed among ancestry-specific effects.

Beyond their genetic characteristics, the loci identified as heterogeneous among the ancestries investigated showed associations with specific phenotypic domains. We observed that certain traits are enriched for associations with variants presenting strong heterogeneity among the ancestry-specific effects. Additionally, GWAS index variants located in independent genomic regions are associated with the same traits. This strongly highlights how cross-ancestry genetic heterogeneity is widespread in the genetic associations with certain phenotypic domains and it does not represent a unique or exceptional event. Across the 14 categories identified within the 843 traits tested (Supplemental Table 1), we observed enrichments for cross-ancestry heterogeneous associations with respect to traits related to blood biomarkers and physical appearance. The enrichment for the first categories showed the strongest significance. Thirty-tree out of the 63 blood biomarkers (52%) showed one or more associations with loci with cross-ancestry heterogeneous effects. These included blood biomarkers related to lipid metabolism (e.g., LDL cholesterol, lipoprotein A, and apolipoprotein B), liver function (alanine aminotransferase and alkaline phosphatase), inflammation (C-reactive protein), glucose metabolism (glucose and glycated haemoglobin), immune function (e.g., eosinophil percentage, monocyte count, and neutrophil count), hematic parameters (e.g., platelet count and erythrocyte distribution width), hormonal regulation (sex hormone-binding globulin), and vitamin levels (vitamin D). Accordingly, we hypothesize that cross-ancestry genetic heterogeneity is not related to a specific function, but rather pervasive across the genetics of multiple molecular phenotypes. Genetic ancestry has been previously associated with the variability of certain blood biomarkers among worldwide populations (48). In line with these genetic differences, multiple studies highlighted that blood biomarkers related to different domains (e.g., cardiometabolic risk or brain aging) present significant differences among ancestry groups (49, 50). These previous data support that some of the observed population differences in disease prevalence have a biological basis and that reference intervals for those biomarkers should be tailored to ancestry. Our present findings not only expand this previous evidence, but also provide novel insights into the underlying mechanisms responsible for this variability. With respect to Δ_LD_ and Δ_LD_ contribution to the genetic heterogeneity of blood biomarkers, we hypothesize that natural selection may have also contributed in addition to the human demographic history. Indeed, loci associated with certain blood biomarkers (e.g., those related to immune function) present genomic selective signatures due to the selective pressures of pathogens, diet changes, and several other environmental variables that shaped human evolutionary history (51). With respect to the second phenotypic domain enriched for loci presenting cross-ancestry heterogeneity, we expect that evolutionary pressures had played a much larger role. Indeed, the traits identified were related to hair and skin appearance. “Hair color (natural, before greying)” showed strong heterogeneity among AMR, CSA, EUR, and MID groups. In line with minimal hair color variation within AFR and EAS individuals, no effect was observed with respect to these human groups. Conversely, alleles associated with black and dark brown hair color showed concordant but heterogeneous effects among the remaining ancestries available in the UKB cohort. Among the seven GWAS index variants located in independent genomic regions (four for black hair color and three for dark brown hair color), the associated allele generally showed the largest effect size with respect to the EUR sample. However, in one case (the association of *NPLOC4* rs9895741*G allele with dark brown hair color), we observed a discordant effect between EUR (beta=0.104, p=2.32×10^−75^) and CSA (beta=-0.194, p=1.95×10^−5^). The phenotype related to skin pigmentation (i.e., “Ease of skin tanning” ranging from “Never tan, only burn” to “Get very tanned”) showed the same pattern across three associated independent GWAS index variants (*TYR* rs7941686*A on chromosome 11, *SLC24A5* rs1426654*G on chromosome 15, and *TCF25* rs577053706*C on chromosome 16): negative GWS association in EUR, consistent effect direction among the other non-AFR samples, and positive effect direction in AFR. The heterogeneity observed within these loci is likely to be affected by the evolutionary mechanisms that shaped human skin and hair pigmentation (52). Indeed, the variability of human pigmentation reflects the subsequent adaptive processes from the protective, dark, eumelanin-enriched coloration useful to protect the naked skin in the tropical regions where *Homo sapiens* originated to the loss of melanin pigmentation that occurred during the dispersal of *Homo sapiens* into non-tropical landmasses (52-54). With respect to this previous knowledge, we expand the understanding of the genetic mechanisms underlying this phenotypic variation, also putting this in the context of the genetics of the human phenotypic spectrum.

Although our investigation provides the first comprehensive evidence regarding how genetic variation across human populations affects the predisposition to complex traits, there are three main limitations to acknowledge. The first two were due to the strong ancestry imbalance among UKB participants (>90% British of European descent). This strongly limited our ability to expand our analyses with respect to this important topic. In particular, our results provide information only regarding differences between European populations and other ancestry groups and the loci and phenotypic domains identified are those with very large heterogeneity. A better representation of human genetic variation would have permitted us to detect heterogeneous signals across non-European populations and to increase our power to detect loci with a lower degree of heterogeneity. Due to the same reason, we limited our investigation to single loci and did not attempt to model cross-ancestry heterogeneity in the context of polygenicity. More diverse genome-wide datasets will support the investigation of genetic heterogeneity with respect to how the polygenic architecture of complex traits varies across human populations. Another important limitation is due to the fact that our analysis was conducted only on the UKB cohort, because this is the only large-scale resource publicly available at this time. Comparing cross-ancestry heterogeneity generated from independent cohorts will permit us to generalize our findings and to understand how the sample characteristics and the recruitment strategies affect gene discovery of complex traits across worldwide populations In conclusion, this study provided novel evidence regarding the predisposition to complex traits in the context of human genetic variation. We observed that loci with heterogeneous effects across ancestries are enriched for traits shaped by human demographic history and natural selection. Both inter-population differences in allele frequencies and LD tagging of regulatory elements affect the genotype-phenotype associations both qualitatively and quantitatively (effect direction and effect size, respectively). However, we showed how the strongest genetic heterogeneity (i.e., discordant effect direction) was mainly driven by differences between European and African populations, while loci with heterogeneous but concordant effects were mainly affected by differences of European ancestry with respect to populations with a complex genetic makeup (e.g., AMR and MID). Finally, although our data contribute to increasing our knowledge regarding how cross-ancestry genetic diversity affect the predisposition to complex traits, they strongly highlight that there is an urgent need for greater population diversity in genome-wide studies.

## Data Availability

Data supporting the findings of this study are available within this article and its additional files.

## ACKNOWLEDGMENTS

We thank the participants and investigators of the Pan-UK Biobank and Dr. Riccardo Pennacchi for the computational support. Yale investigators acknowledges support from the National Institutes of Health via R21 DA047527, R21 DC018098, and F32 MH122058. The sources of funding had no role in the design of the study and collection, analysis, and interpretation of data and in writing the manuscript.

## CONFLICT OF INTEREST

The authors reported no biomedical financial interests or potential conflicts of interest.

## Notes

### Competing Interest Statement

The authors have declared no competing interest.

### Funding Statement

Yale investigators acknowledge support from the National Institutes of Health via R21 DA047527, R21 DC018098, and F32 MH122058. The sources of funding had no role in the design of the study and collection, analysis, and interpretation of data and in writing the manuscript.

### Author Declarations

This study was conducted using summary association data generated by previous studies. Owing to the use of previously collected, deidentified, aggregated data, this study did not require institutional review board approval.

## References

1 Visscher, P.M., Wray, N.R., Zhang, Q., Sklar, P., McCarthy, M.I., Brown, M.A. and Yang, J. (2017) 10 Years of GWAS Discovery: Biology, Function, and Translation. Am J Hum Genet, 101, 5–22.

2 Buniello, A., MacArthur, J.A.L., Cerezo, M., Harris, L.W., Hayhurst, J., Malangone, C., McMahon, A., Morales, J., Mountjoy, E., Sollis, E. et al. (2019) The NHGRI-EBI GWAS Catalog of published genome-wide association studies, targeted arrays and summary statistics 2019. Nucleic Acids Res, 47, D1005–D1012.

3 Sullivan, P.F., Agrawal, A., Bulik, C.M., Andreassen, O.A., Borglum, A.D., Breen, G., Cichon, S., Edenberg, H.J., Faraone, S.V., Gelernter, J. et al. (2018) Psychiatric Genomics: An Update and an Agenda. Am J Psychiatry, 175, 15–27.

4 Thompson, P.M., Stein, J.L., Medland, S.E., Hibar, D.P., Vasquez, A.A., Renteria, M.E., Toro, R., Jahanshad, N., Schumann, G., Franke, B. et al. (2014) The ENIGMA Consortium: large-scale collaborative analyses of neuroimaging and genetic data. Brain Imaging Behav, 8, 153–182.

5 Kim, Y., Han, B.G. and Ko, G.E.S.g. (2017) Cohort Profile: The Korean Genome and Epidemiology Study (KoGES) Consortium. Int J Epidemiol, 46, e20.

6 Colodro-Conde, L., Cross, S.M., Lind, P.A., Painter, J.N., Gunst, A., Jern, P., Johansson, A., Lund Maegbaek, M., Munk-Olsen, T., Nyholt, D.R. et al. (2017) Cohort Profile: Nausea and vomiting during pregnancy genetics consortium (NVP Genetics Consortium). Int J Epidemiol, 46, e17.

7 Fan, C.T., Lin, J.C. and Lee, C.H. (2008) Taiwan Biobank: a project aiming to aid Taiwan’s transition into a biomedical island. Pharmacogenomics, 9, 235–246.

8 Kubo, M. and Guest, E. (2017) BioBank Japan project: Epidemiological study. J Epidemiol, 27, S1.

9 Sudlow, C., Gallacher, J., Allen, N., Beral, V., Burton, P., Danesh, J., Downey, P., Elliott, P., Green, J., Landray, M. et al. (2015) UK biobank: an open access resource for identifying the causes of a wide range of complex diseases of middle and old age. PLoS Med, 12, e1001779.

10 Check Hayden, E. (2017) The rise and fall and rise again of 23andMe. Nature, 550, 174–177.

11 Evangelou, E., Warren, H.R., Mosen-Ansorena, D., Mifsud, B., Pazoki, R., Gao, H., Ntritsos, G., Dimou, N., Cabrera, C.P., Karaman, I. et al. (2018) Genetic analysis of over 1 million people identifies 535 new loci associated with blood pressure traits. Nat Genet, 50, 1412–1425.

12 Timmers, P.R., Mounier, N., Lall, K., Fischer, K., Ning, Z., Feng, X., Bretherick, A.D., Clark, D.W., e, Q.C., Agbessi, M. et al. (2019) Genomics of 1 million parent lifespans implicates novel pathways and common diseases and distinguishes survival chances. Elife, 8.

13 Karlsson Linner, R., Biroli, P., Kong, E., Meddens, S.F.W., Wedow, R., Fontana, M.A., Lebreton, M., Tino, S.P., Abdellaoui, A., Hammerschlag, A.R. et al. (2019) Genome-wide association analyses of risk tolerance and risky behaviors in over 1 million individuals identify hundreds of loci and shared genetic influences. Nat Genet, 51, 245–257.

14 Lee, J.J., Wedow, R., Okbay, A., Kong, E., Maghzian, O., Zacher, M., Nguyen-Viet, T.A., Bowers, P., Sidorenko, J., Karlsson Linner, R. et al. (2018) Gene discovery and polygenic prediction from a genome-wide association study of educational attainment in 1.1 million individuals. Nat Genet, 50, 1112–1121.

15 Weigl, K., Chang-Claude, J., Knebel, P., Hsu, L., Hoffmeister, M. and Brenner, H. (2018) Strongly enhanced colorectal cancer risk stratification by combining family history and genetic risk score. Clin Epidemiol, 10, 143–152.

16 Sparano, J.A., Gray, R.J., Ravdin, P.M., Makower, D.F., Pritchard, K.I., Albain, K.S., Hayes, D.F., Geyer, C.E., Jr., Dees, E.C., Goetz, M.P. et al. (2019) Clinical and Genomic Risk to Guide the Use of Adjuvant Therapy for Breast Cancer. N Engl J Med, 380, 2395–2405.

17 Khera, A.V., Chaffin, M., Wade, K.H., Zahid, S., Brancale, J., Xia, R., Distefano, M., Senol-Cosar, O., Haas, M.E., Bick, A. et al. (2019) Polygenic Prediction of Weight and Obesity Trajectories from Birth to Adulthood. Cell, 177, 587–596 e589.

18 Inouye, M., Abraham, G., Nelson, C.P., Wood, A.M., Sweeting, M.J., Dudbridge, F., Lai, F.Y., Kaptoge, S., Brozynska, M., Wang, T. et al. (2018) Genomic Risk Prediction of Coronary Artery Disease in 480,000 Adults: Implications for Primary Prevention. J Am Coll Cardiol, 72, 1883–1893.

19 Sirugo, G., Williams, S.M. and Tishkoff, S.A. (2019) The Missing Diversity in Human Genetic Studies. Cell, 177, 1080.

20 Martin, A.R., Kanai, M., Kamatani, Y., Okada, Y., Neale, B.M. and Daly, M.J. (2019) Clinical use of current polygenic risk scores may exacerbate health disparities. Nat Genet, 51, 584–591.

21 Mostafavi, H., Harpak, A., Conley, D., Pritchard, J.K. and Przeworski, M. (2019) Variable prediction accuracy of polygenic scores within an ancestry group. bioRxiv, in press., 629949.

22 Martin, A.R., Gignoux, C.R., Walters, R.K., Wojcik, G.L., Neale, B.M., Gravel, S., Daly, M.J., Bustamante, C.D. and Kenny, E.E. (2017) Human Demographic History Impacts Genetic Risk Prediction across Diverse Populations. Am J Hum Genet, 100, 635–649.

23 Duncan, L., Shen, H., Gelaye, B., Meijsen, J., Ressler, K., Feldman, M., Peterson, R. and Domingue, B. (2019) Analysis of polygenic risk score usage and performance in diverse human populations. Nat Commun, 10, 3328.

24 Gaziano, J.M., Concato, J., Brophy, M., Fiore, L., Pyarajan, S., Breeling, J., Whitbourne, S., Deen, J., Shannon, C., Humphries, D. et al. (2016) Million Veteran Program: A mega-biobank to study genetic influences on health and disease. J Clin Epidemiol, 70, 214–223.

25 Sankar, P.L. and Parker, L.S. (2017) The Precision Medicine Initiative’s All of Us Research Program: an agenda for research on its ethical, legal, and social issues. Genet Med, 19, 743–750.

26 Daub, J.T., Hofer, T., Cutivet, E., Dupanloup, I., Quintana-Murci, L., Robinson-Rechavi, M. and Excoffier, L. (2013) Evidence for polygenic adaptation to pathogens in the human genome. Mol Biol Evol, 30, 1544–1558.

27 Hofer, T., Ray, N., Wegmann, D. and Excoffier, L. (2009) Large allele frequency differences between human continental groups are more likely to have occurred by drift during range expansions than by selection. Ann Hum Genet, 73, 95–108.

28 Iorio, A., De Angelis, F., Di Girolamo, M., Luigetti, M., Pradotto, L.G., Mazzeo, A., Frusconi, S., My, F., Manfellotto, D., Fuciarelli, M. et al. (2017) Population diversity of the genetically determined TTR expression in human tissues and its implications in TTR amyloidosis. BMC Genomics, 18, 254.

29 Polimanti, R., Yang, C., Zhao, H. and Gelernter, J. (2015) Dissecting ancestry genomic background in substance dependence genome-wide association studies. Pharmacogenomics, 16, 1487–1498.

30 Bycroft, C., Freeman, C., Petkova, D., Band, G., Elliott, L.T., Sharp, K., Motyer, A., Vukcevic, D., Delaneau, O., O’Connell, J. et al. (2018) The UK Biobank resource with deep phenotyping and genomic data. Nature, 562, 203–209.

31 1000 Genomes Project Consortium, Auton, A., Brooks, L.D., Durbin, R.M., Garrison, E.P., Kang, H.M., Korbel, J.O., Marchini, J.L., McCarthy, S., McVean, G.A. et al. (2015) A global reference for human genetic variation. Nature, 526, 68–74.

32 Li, J.Z., Absher, D.M., Tang, H., Southwick, A.M., Casto, A.M., Ramachandran, S., Cann, H.M., Barsh, G.S., Feldman, M., Cavalli-Sforza, L.L. et al. (2008) Worldwide human relationships inferred from genome-wide patterns of variation. Science, 319, 1100–1104.

33 Zhou, W., Nielsen, J.B., Fritsche, L.G., Dey, R., Gabrielsen, M.E., Wolford, B.N., LeFaive, J., VandeHaar, P., Gagliano, S.A., Gifford, A. et al. (2018) Efficiently controlling for case-control imbalance and sample relatedness in large-scale genetic association studies. Nat Genet, 50, 1335–1341.

34 Boyle, A.P., Hong, E.L., Hariharan, M., Cheng, Y., Schaub, M.A., Kasowski, M., Karczewski, K.J., Park, J., Hitz, B.C., Weng, S. et al. (2012) Annotation of functional variation in personal genomes using RegulomeDB. Genome Res, 22, 1790–1797.

35 Machiela, M.J. and Chanock, S.J. (2015) LDlink: a web-based application for exploring population-specific haplotype structure and linking correlated alleles of possible functional variants. Bioinformatics, 31, 3555–3557.

36 Machiela, M.J. and Chanock, S.J. (2018) LDassoc: an online tool for interactively exploring genome-wide association study results and prioritizing variants for functional investigation. Bioinformatics, 34, 887–889.

37 Wojcik, G.L., Graff, M., Nishimura, K.K., Tao, R., Haessler, J., Gignoux, C.R., Highland, H.M., Patel, Y.M., Sorokin, E.P., Avery, C.L. et al. (2019) Genetic analyses of diverse populations improves discovery for complex traits. Nature, 570, 514–518.

38 Magi, R., Horikoshi, M., Sofer, T., Mahajan, A., Kitajima, H., Franceschini, N., McCarthy, M.I., Cogent-Kidney Consortium, T.D.G.C. and Morris, A.P. (2017) Trans-ethnic meta-regression of genome-wide association studies accounting for ancestry increases power for discovery and improves fine-mapping resolution. Hum Mol Genet, 26, 3639–3650.

39 Asimit, J.L., Hatzikotoulas, K., McCarthy, M., Morris, A.P. and Zeggini, E. (2016) Trans-ethnic study design approaches for fine-mapping. Eur J Hum Genet, 24, 1330–1336.

40 Koyama, S., Ito, K., Terao, C., Akiyama, M., Horikoshi, M., Momozawa, Y., Matsunaga, H., Ieki, H., Ozaki, K., Onouchi, Y. et al. (2020) Population-specific and trans-ancestry genome-wide analyses identify distinct and shared genetic risk loci for coronary artery disease. Nat Genet, 52, 1169–1177.

41 Gelernter, J., Sun, N., Polimanti, R., Pietrzak, R., Levey, D.F., Bryois, J., Lu, Q., Hu, Y., Li, B., Radhakrishnan, K. et al. (2019) Genome-wide association study of post-traumatic stress disorder reexperiencing symptoms in >165,000 US veterans. Nat Neurosci, 22, 1394–1401.

42 Mahajan, A., Spracklen, C.N., Zhang, W., Ng, M.C.Y., Petty, L.E., Kitajima, H., Yu, G.Z., Rueger, S., Speidel, L., Kim, Y.J. et al. (2020) Trans-ancestry genetic study of type 2 diabetes highlights the power of diverse populations for discovery and translation. medRxiv, in press., 2020.2009.2022.20198937.

43 Scerri, E.M.L., Chikhi, L. and Thomas, M.G. (2019) Beyond multiregional and simple out-of-Africa models of human evolution. Nat Ecol Evol, 3, 1370–1372.

44 Groucutt, H.S., Petraglia, M.D., Bailey, G., Scerri, E.M., Parton, A., Clark-Balzan, L., Jennings, R.P., Lewis, L., Blinkhorn, J., Drake, N.A. et al. (2015) Rethinking the dispersal of Homo sapiens out of Africa. Evol Anthropol, 24, 149–164.

45 Bentley, A.R., Callier, S.L. and Rotimi, C.N. (2020) Evaluating the promise of inclusion of African ancestry populations in genomics. NPJ Genom Med, 5, 5.

46 Gravel, S., Zakharia, F., Moreno-Estrada, A., Byrnes, J.K., Muzzio, M., Rodriguez-Flores, J.L., Kenny, E.E., Gignoux, C.R., Maples, B.K., Guiblet, W. et al. (2013) Reconstructing Native American migrations from whole-genome and whole-exome data. PLoS Genet, 9, e1004023.

47 Scott, E.M., Halees, A., Itan, Y., Spencer, E.G., He, Y., Azab, M.A., Gabriel, S.B., Belkadi, A., Boisson, B., Abel, L. et al. (2016) Characterization of Greater Middle Eastern genetic variation for enhanced disease gene discovery. Nat Genet, 48, 1071–1076.

48 Sjaarda, J., Gerstein, H.C., Kutalik, Z., Mohammadi-Shemirani, P., Pigeyre, M., Hess, S. and Pare, G. (2020) Influence of Genetic Ancestry on Human Serum Proteome. Am J Hum Genet, 106, 303–314.

49 Morris, J.C., Schindler, S.E., McCue, L.M., Moulder, K.L., Benzinger, T.L.S., Cruchaga, C., Fagan, A.M., Grant, E., Gordon, B.A., Holtzman, D.M. et al. (2019) Assessment of Racial Disparities in Biomarkers for Alzheimer Disease. JAMA Neurol, 76, 264–273.

50 Hackler, E., 3rd, Lew, J. Gore, M.O., Ayers, C.R., Atzler, D., Khera, A., Rohatgi, A., Lewis, A., Neeland, I., Omland, T. et al. (2019) Racial Differences in Cardiovascular Biomarkers in the General Population. J Am Heart Assoc, 8, e012729.

51 Quintana-Murci, L. (2019) Human Immunology through the Lens of Evolutionary Genetics. Cell, 177, 184–199.

52 Pavan, W.J. and Sturm, R.A. (2019) The Genetics of Human Skin and Hair Pigmentation. Annu Rev Genomics Hum Genet, 20, 41–72.

53 Jablonski, N.G. and Chaplin, G. (2017) The colours of humanity: the evolution of pigmentation in the human lineage. Philos Trans R Soc Lond B Biol Sci, 372.

54 Wilde, S., Timpson, A., Kirsanow, K., Kaiser, E., Kayser, M., Unterlander, M., Hollfelder, N., Potekhina, I.D., Schier, W., Thomas, M.G. et al. (2014) Direct evidence for positive selection of skin, hair, and eye pigmentation in Europeans during the last 5,000 y. Proc Natl Acad Sci U S A, 111, 4832–4837.

